# The UK experience of stereoelectroencephalography in children: An analysis of factors predicting the identification of a seizure onset zone and subsequent seizure freedom

**DOI:** 10.1101/2021.03.05.21252613

**Authors:** Children’s Epilepsy Surgery Service, Aswin Chari

## Abstract

**Importance:** Stereoelectroencephalography (SEEG) is more frequently being used in the pre-surgical evaluation of children with focal epilepsy. Many factors affect the rate of identification of a definable seizure onset zone (SOZ) and subsequent seizure freedom following SEEG-guided epilepsy surgery, which have not been systematically examined in multi-centre studies.

**Objectives:** Determine the rates and factors that predict (a) whether or not a definable putative SOZ was identified on SEEG and (b) subsequent seizure freedom following surgical intervention.

**Design:** Retrospective cohort study

**Setting:** Multicentre study involving 6 of 7 UK Children’s Epilepsy Surgery Service centres that perform paediatric SEEG in the UK.

**Participants:** All children undergoing SEEG from 2014 - March 2019 were included. Demographic, non-invasive evaluation, SEEG and operative factors were collected retrospectively from patient records.

**Main Outcomes:** The two main outcome measures were (a) whether or not a definable putative SOZ was identified on SEEG (binary yes/no outcome) and (b) subsequent seizure freedom following surgical intervention (Engel classification)

**Findings:** One hundred and thirty-five patients underwent 139 SEEG explorations using a total of 1767 electrodes. A definable SOZ was identified in 117 patients (85.7%); odds of successfully finding a SOZ were 6.4x greater for non-motor seizures compared to motor seizures (p=0.02) and 3.6x more if ≥ 4 seizures were recorded during SEEG (p=0.03). Of 100 patients undergoing surgical treatment, 47 (47.0%) had an Engel class I outcome at a median follow-up of 1.3 years; the only factor associated with outcome was indication for SEEG (p=0.03). SEEG was safe with one (0.7%) haematoma requiring surgical evacuation and no long-term neurological deficits as a result of the procedure.

**Conclusions and Relevance:** This large nationally representative cohort illustrates that, in these patients who may not have otherwise been offered resective surgery, SEEG-guided surgery can still achieve high rates of seizure freedom. Seizure semiology and the number of seizures recorded during SEEG are important factors in the identification of a definable SOZ and the indication for SEEG is an important factor in post-operative outcomes.

## Introduction

Surgery for refractory focal epilepsy in children is effective, with around 70% becoming seizure free (Engel Class I) following resective surgery in carefully selected candidates.^1^ Seizure freedom improves quality of life and, therefore, increasing numbers of children now undergo presurgical evaluation. More complex cases being considered, including those without clear radiological abnormalities or in whom there is uncertain localisation in the non-invasive studies.^1,2^ The increased complexity has resulted in more frequent use of invasive electroencephalography (EEG), particularly stereoelectroencephalography (SEEG) as it provides better topographic accuracy, the ability to explore spatially distant and deep areas (including bilateral and insular implantations) and therapeutic options during the monitoring such as radiofrequency thermocoagulation. ^1,3^

Recent studies have shown that advances in imaging, planning and robotic-assisted surgery have made SEEG a safe tool in children, with low rates of adverse events such as haemorrhage (particularly symptomatic intracranial haemorrhage causing permanent neurological deficit) and infection. ^3–13^ Despite these advances, the rate of seizure free outcomes following epilepsy surgery has remained relatively static over time, at around 70% and this proportion is even lower following SEEG-guided resective surgery, at 50-67%.^4,9,13–15^ This may be attributable, at least in part, to the selection of more complex candidates who may not have been considered for surgery in the past. In the context of SEEG, the definition of ‘success’ is in itself a complex consideration as it may be variably interpreted as identification of the seizure onset zone (SOZ), offering subsequent surgery or via the more traditional surgical outcomes of seizure freedom or improved quality of life. The rates of each of these measures may vary as they are dependent on a number of factors including the selection of candidates for SEEG, the implantation plan, subsequent interpretation of the SEEG recordings to devise a surgical strategy as well as the adequacy of the operation itself (Figure 1). All of these may be influenced by institutional ethos and the biases of the multidisciplinary presurgical evaluation teams.

**Figure 1.**
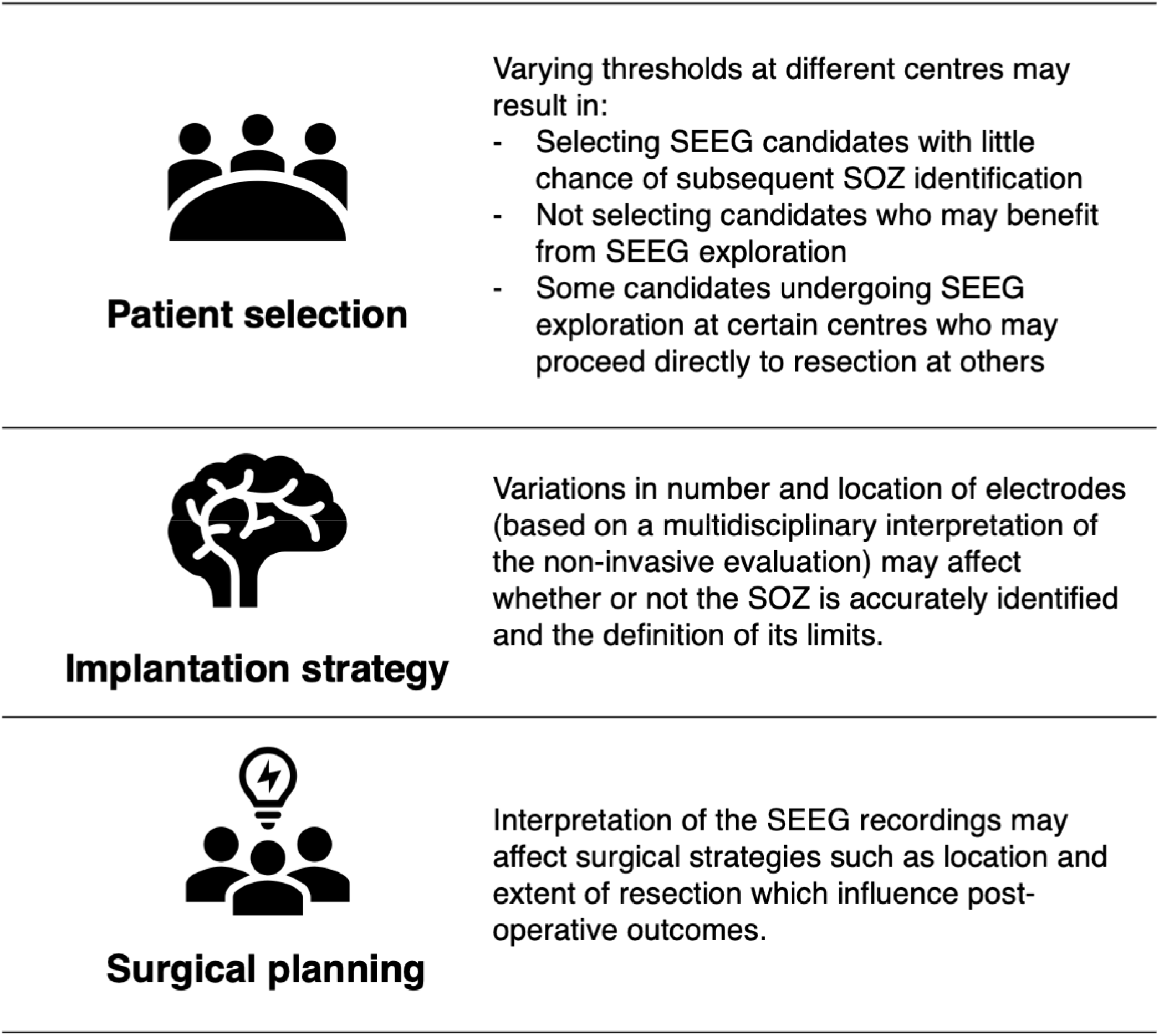
Schematic illustration of the factors affecting surgical success in an SEEG programme. SEEG = stereoelectroencephalography, SOZ = seizure onset zone.

In order to explore these factors in a real-world setting, we sought to undertake a nationwide multi-centre study of the United Kingdom (UK) experience of paediatric SEEG. The two specific aims were to analyse preoperative and SEEG factors that predicted (a) whether or not a definable putative SOZ was identified on SEEG and (b) subsequent seizure freedom following surgical intervention.

## Methods

### Design

Multi-centre retrospective cohort observational study. This manuscript has been reported in accordance with the STROBE Guidelines (available in the Supplementary Material).^16^

### Centres

All centres performing paediatric epilepsy surgery and SEEG in England are part of a centrally commissioned NHS England Children’s Epilepsy Surgery Service (CESS), initiated in 2012. All of these centres and the single centre performing paediatric SEEG in Scotland were invited to participate in this retrospective cohort study, encompassing all centres performing paediatric SEEG in the UK.

Six of the seven centres agreed to participate, namely Great Ormond Street Hospital (GOSH, London), King’s College Partners (KCP, London), Bristol Royal Hospital for Children (Bristol), Manchester Children’s Hospital (Manchester), Alder Hey Children’s Hospital (Liverpool) and Royal Hospital for Sick Children (Edinburgh). This study was approved by the Clinical Audit Department at GOSH (registration #1870). In addition, each centre registered the study as a retrospective service evaluation with their local research and development office.

### Case Selection

All children who underwent SEEG at these paediatric centres between 2014 and the end of March 2019 were eligible for inclusion. There were no exclusion criteria. Patients were selected for SEEG based on local epilepsy multidisciplinary team (MDT) decision following non-invasive evaluation that included at least an epilepsy-protocol MRI scan (defined locally at each institution), EEG video-telemetry and neurodevelopmental/neuropsychological evaluation. Other adjunctive investigations may have included positron emission tomography (PET), magnetoencephalography (MEG), functional MRI (fMRI) and ictal single-photon emission computed tomography (SPECT) scans, at the discretion of the local MDT. The CESS network also conducts a national MDT meeting that allows complex cases to be discussed; although this ensures some alignment of decision making across the country, the final decision and implantation strategies remain at the discretion of the local team at each centre.

### Data Collection

Data were collected from patient records via a piloted proforma between September 2019 and December 2020. Data were collected in a number of domains, detailed in Supplementary Table 1. To reduce bias, the majority of the data was designed to be readily available in the presurgical MDT proforma, which is largely similar across the centres. The two outcome measures of interest were (a) a binary outcome of whether or not a definable SOZ was identified following the SEEG exploration and (b) the post-operative Engel class at last follow-up that was also dichotomised into class I (seizure free) and class II-IV (not seizure free).

### Statistical analysis

The statistical analysis was conducted according to a pre-specified analysis plan incorporating demographic, pre-surgical evaluation, SEEG (and, for the 2^nd^ analysis, resective operation) factors into a stepwise binary logistic regression model to identify factors that predicted (a) the identification of a SOZ on SEEG and (b) subsequent seizure freedom following resection. For the 2 regression analyses performed, only the 2^nd^ exploration was taken into consideration for patients who had undergone 2 explorations. Cases with missing data would have been excluded but all records were complete.

In addition, a number of other descriptive analyses were performed, which were explored as they were thought to be of clinical interest or were deemed to warrant further exploration given the results of the pre-specified statistical analyses. All statistical analyses were performed on Microsoft Excel v16 (©Microsoft Inc), SPSS v24 (©IBM Inc) and Matlab R2018b (©The Mathworks Inc). P-values <0.05 were considered statistically significant.

## Results

### Demographics

A total of 139 SEEG explorations were conducted in 135 patients across the 6 centres during the inclusion period. The number of SEEG cases increased with time across all the centres (Figure 2a). The median age at SEEG was 11 years (range 3-19) with a bimodal distribution (peaks around ages 9 and 16) and the median duration of epilepsy at SEEG implantation was 7 years (range 0-19 years) (Figure 2b). The most common indications for SEEG (classified in Supplementary Table 1) were ‘lesion positive, define extent of lesion’ (29.5%) and ‘lesion negative’ (28.1%) (Figure 2c); of the 4 repeat explorations, 3 were MRI lesion negative cases.

**Figure 2.**
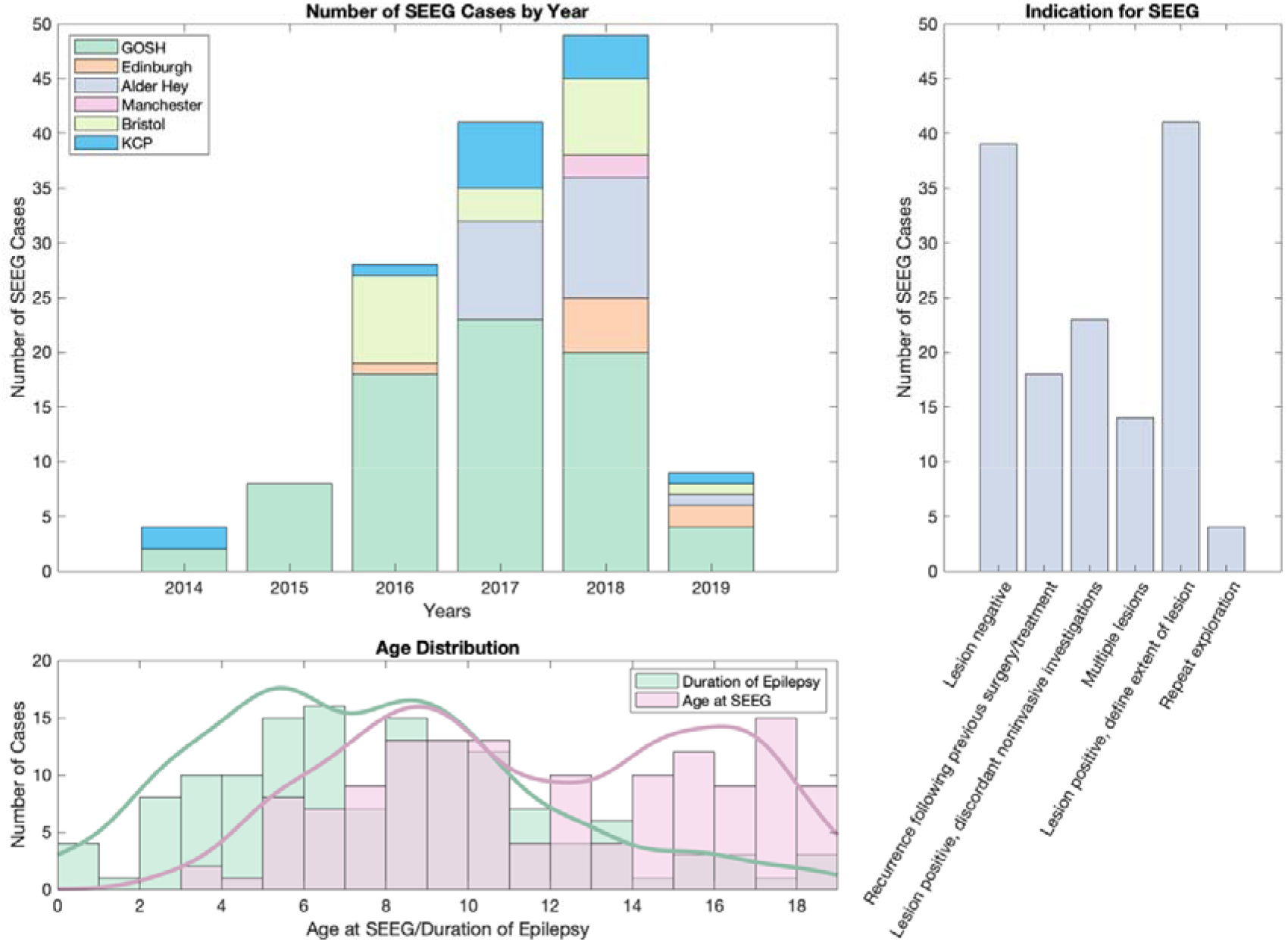
Demographics of the multi-centre SEEG cohort. (a) Number of SEEG cases split by year and centre. Note that 2019 only encompasses the first 3 months of the year. (b) Histograms and kernel curves of distribution of age for SEEG cases (pink) and duration of epilepsy (green). (c) Indication for SEEG.

### Non-Invasive Evaluation

Prior to invasive evaluation, all patients underwent detailed clinical evaluation, MRI scans and scalp EEG-video telemetry (Supplementary Table 2). Seventeen (17.0) percent had pre-existing focal neurological deficit on examination, 31.1% had significant neuropsychological impairment (FSIQ < 70) and 14.8% had a neuropsychiatric diagnosis (eg autism, anxiety, depression). At the discretion of the local team, a proportion of patients underwent additional investigations: interictal PET (57.8%), MEG (18.5%), ictal SPECT (17.8%) and language fMRI (22.2%).

### SEEG Implantation & Surgical Safety

Apart from the first 6 cases at GOSH and the first case in Edinburgh (frameless neuronavigated procedures) and the first 2 cases at KCP (frame-based arc procedures), all cases were performed using a frame-based robotic-assisted technique (Renishaw Neuromate system), which is detailed elsewhere.^8^ A total of 1767 electrodes were implanted across the 139 explorations, with a median [IQR] of 12 [10-15] electrodes per implantation (Figure 3a). Dividing the brain into 10 lobes (frontal, temporal, parietal, occipital and insula in each hemisphere), a median [IQR] of 4 [3-4] lobes were explored (Figure 3a). A ratio of electrodes/lobe was calculated as a surrogate marker of confidence in the implantation hypothesis, with high ratios indicating increased confidence (Figure 3b). There were significant differences between the ratios for each indication (Kruskal-Wallis test, p=9.3×10^−6^, Figure 3d). There were 98 (70.5%) unilateral and 41 (29.5%) bilateral explorations; the left hemisphere lobes were explored more frequently than the right and, the frontal and temporal lobes were explored more than other areas (Figure 3c).

**Figure 3.**
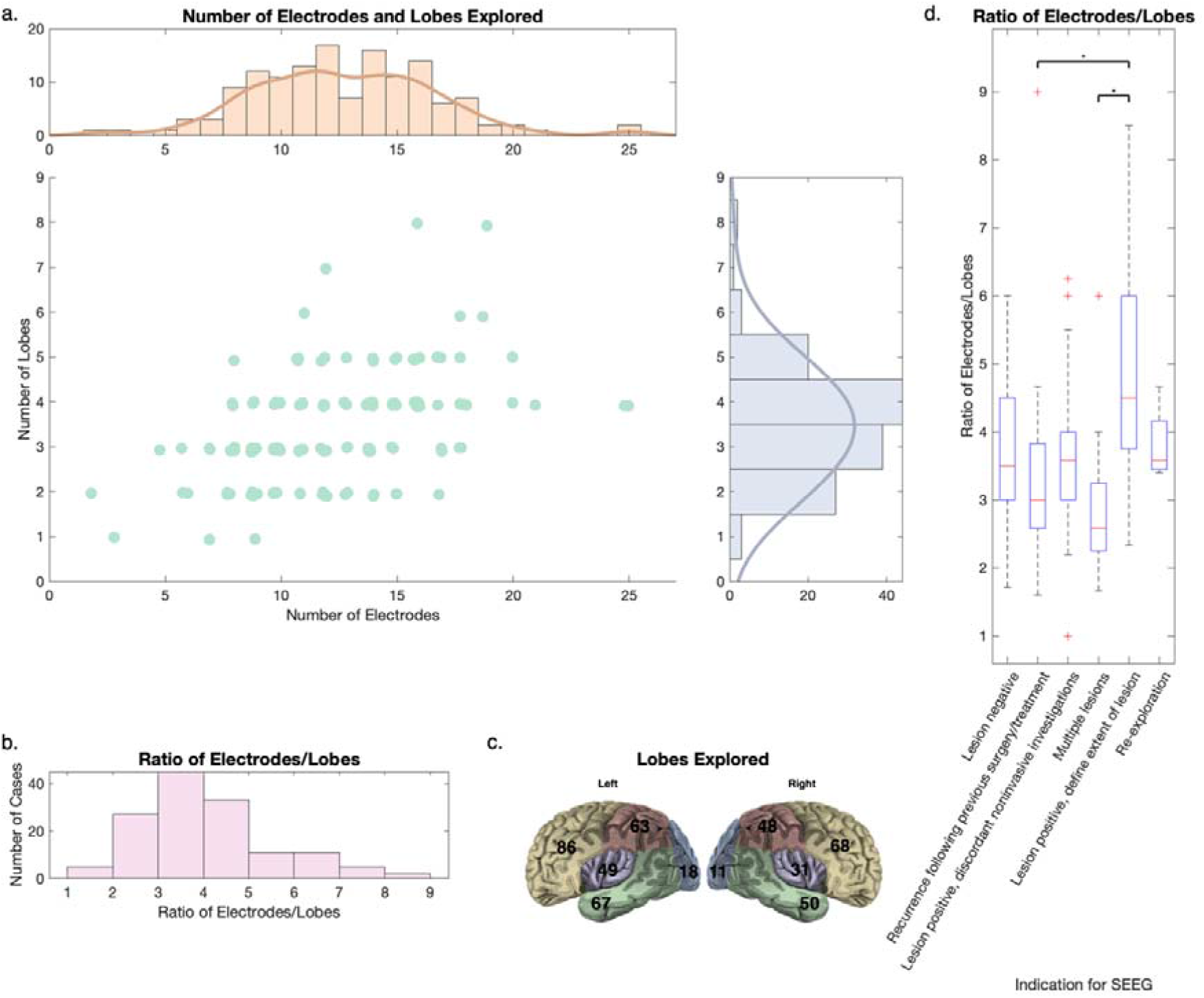
SEEG exploration factors in 139 explorations. (a) Scatter plot of number of electrodes and lobes explored with histograms for each shown separately at the ends of the axes, indicating a range in both domains. The moderate correlation between the two (Spearman correlation r = 0.46, p=1×10^−8^) indicates that it is not necessarily that more lobes equate to more electrodes. (b) Ratio of electrodes/lobes, a novel surrogate indicator of ‘confidence’ in the pre-implantation hypothesis as a more limited spatial exploration (higher ratio) is likely to indicate more confidence from the non-invasive investigations. (c) Visual representation of the lobes explored, showing more exploration of the left sided lobes. (d) Box plots showing the ratio of electrodes/lobes by indication for SEEG. Kruskal Wallis testing revealed significant differences between the groups (p=9.3×10^−6^) with post-hoc pairwise comparisons showing significant differences between the ‘lesion positive, discordant non-invasive investigations’ group and ‘recurrence following previous surgery/treatment’ (p=3×10^−4^) and ‘multiple lesions’ (p=6×10^−5^) groups following correction for multiple comparisons using the Tukey method.

Recording occurred for a median [IQR] of 7 [5-7] days, not accounting for 2 cases where there were no recordings (one due to bleeding and one due to electrodes being pulled out by the patient on return to the ward). Stimulation testing was performed in 111 cases (79.9%) with a mixture of 1Hz and 50Hz stimulation to encompass both seizure and functional stimulation; 68 (61.3%) had seizures stimulated, in whom 59 (86.8%) were thought consistent with their habitual seizures.

In terms of safety, only one case (0.7%) had significant bleeding requiring surgical evacuation, three (2.2%) had minor asymptomatic bleeding identified on the routine post-operative CT scan and five (3.6%) had one or more electrodes either malpositioned (extradural) or pulled out – in one case, this resulted in no recordings being gathered. Overall, no long-term neurologic deficits were attributable to the SEEG procedures.

### Identification of a definable SOZ

In order to identify factors that predicted the positive identification of a definable SOZ, we considered only the 2^nd^ exploration in those that were implanted twice (n=4) and excluded those with no recordings (n=2), giving a total of 133 patients. A SOZ was identified in 117 of these (88.0%). Pre-operative and operative variables were assessed for differences between the patients in whom a SOZ was or was not identified (Supplementary Table 3). In the univariate analysis, a MEG scan was less commonly performed (p=0.04), ≥ 4 seizures were more commonly recorded during SEEG (p=0.04), and a habitual seizure was more commonly stimulated (p=0.03) when a SOZ was identified.

A binomial logistic regression model was created using the variables in Supplementary Table 3 that had a p-value of <= 0.25. Backwards elimination resulted in a statistically significant model (p=0.003) with 2 significant variables, namely semiology type (p=0.02) and the number of seizures recorded during SEEG (p=0.03). Odds of successfully finding a SOZ were 6.4x [95% CI 1.3 – 30.2] more for non-motor seizures (compared to motor seizures) and 3.6x [95% CI 1.1 – 11.1] more if ≥ 4 seizures were recorded during SEEG.

### Surgical Resection and Subsequent Seizure Freedom

Overall, 105 patients (78.9% of all patients, 89.7% of those in whom a SOZ was identified) were offered further surgical intervention for their epilepsy (excluding vagal nerve stimulator implantation). The 12 patients who were not offered surgical treatment were due to high risk of deficit due to overlap with functional motor or language areas (6 patients), a widespread SOZ (6 patients) and low seizure burden in the period following SEEG (1 patient). A further 5 patients did not undergo surgical intervention – two transferred to the adult services for their surgery and three opted against proceeding with surgery due to either low seizure burden or high risk of deficit.

The interventions received by the 100 patients and outcomes at last follow-up (median 1.3 years from the last surgical procedure, IQR 1.0-1.9 years, 85.0% with at least 1 year follow-up) are shown in Figure 4a. Overall, 47 patients (47.0% of all those undergoing SEEG-guided treatment or 34.8% of all patients in this series) had an Engel class I outcome.

**Figure 4.**
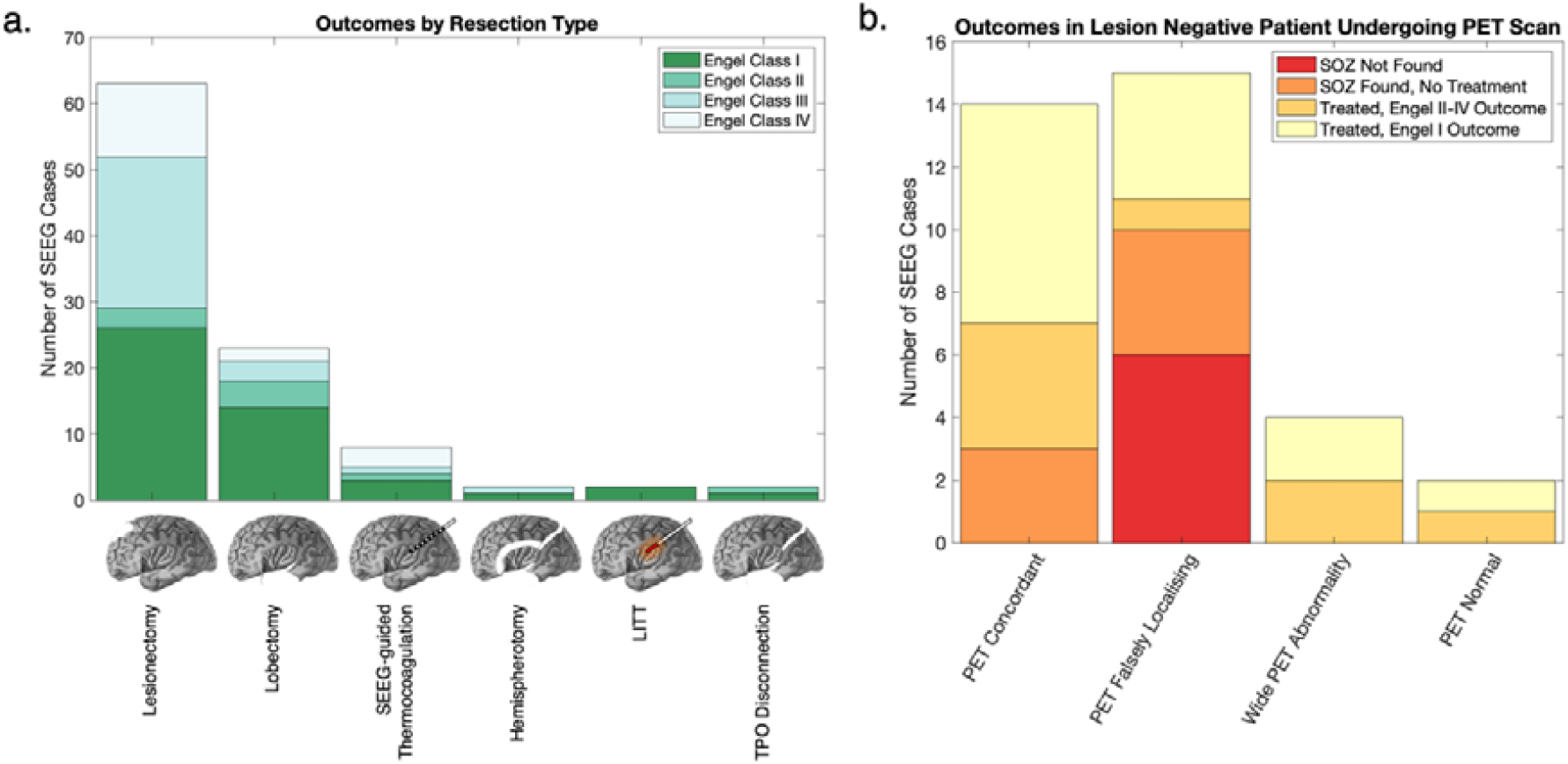
(a) Outcome by resection type. Three patients underwent 2 lesionectomy procedures. Four underwent thermocoagulation prior to other treatment (two lesionectomy, two LITT) and have been classified as their 2^nd^ (definitive) treatment. Lesionectomy involves an SEEG-tailored focal resection of the presumed epileptogenic zone whilst lobectomy involves a larger resection of the lobe. LITT = laser interstitial thermal therapy, TPO disconnection = temporo-parieto-occipital disconnection. (b) Outcomes in lesion negative SEEG cases stratified by finding on the pre-SEEG PET scan. Note that although a lower proportion of those with a falsely localising PET scan went on to have an Engel class I outcome, the proportion of those with an Engel class I outcome as a function of those receiving treatment is similar across groups.

Select SEEG and operative variables were assessed for differences between those that did and did not achieve an Engel class I outcome following resective surgery (n=92; cases undergoing thermocoagulation only were excluded as this is primarily used as a prognostic test rather than definitive treatment across the CESS centres). Overall, 44 (47.8%) patients had an Engel Class I outcome. Significant variables on univariate analysis included the indication for SEEG (p=0.01) and the post-operative histology (p=0.006) (Supplementary Table 4).

A binomial logistic regression model was created using the variables in Supplementary Table 4 that had a p-value of <= 0.25. Backwards elimination resulted in a statistically significant model (p=2×10^−5^) with 2 variables, one of which was statistically significant (indication for SEEG, p=0.03) and one not (histology, p=0.10). Within the indication for SEEG, ‘recurrence following surgery/treatment’ had a 5.9x lower odds of achieving seizure freedom (p=0.002) compared to the ‘lesion negative’ cohort. Within the histology group, those who had a histology of FCD Type 2a and 2b had a 8.9x and 10.4x higher odds of seizure freedom (p=0.02 and p=0.01 respectively) compared to a non-diagnostic/other histology.

### Additional exploratory analyses

As these are additional post-hoc exploratory analyses, numbers and percentages are reported but no statistical tests performed. The particular cohorts were selected because of clinical interest.

### SEEG in children with tuberous sclerosis complex (TSC)

Thirteen patients underwent SEEG in the context of TSC, with a median age of 8 years (range 5-15). A SOZ was identified in 11 of these (84.6%) and all underwent resective surgery which involved a single tuber (3 patients), multiple tubers (6 patients) or multiple tubers + mesial temporal structures (2 patients). Engel class I was achieved in 1 patient (7.7% of all patients explored), class II in 2 (15.4%), class III in 6 (46.2%) and class IV in 2 (15.4%).

### Re-explorations following previous intervention

Of the 20 such cases, 17 had undergone resections (with or without electrocorticography guidance), 3 had undergone disconnective procedures (2 temporo-parieto-occipital (TPO) disconnections and a corpus callosotomy) and one had undergone gamma knife radiosurgery (to a nodular heterotopia). Histologies from the resective/TPO procedures were focal cortical dysplasia (FCD) type IIa (5/18), FCD type IIb (4/18), non-diagnostic/other (8/18) and FCD type 1 (1/18). The median duration (range) from first operation to SEEG was 4 (1-13) years.

A SOZ was identified in 15 (75.0%) patients, of whom 10 underwent subsequent further resective/disconnective surgery and 2 underwent radiofrequency thermocoagulation. Overall, Engel class I was achieved in 2 patients (10% of all patients explored), class II in 3 (15.0%), class III in 6 (30.0%) and class IV in 1 (5.0%). Interestingly, of the 8 patients that underwent a repeat lesionectomy, none achieved a class I outcome.

### The Utility of PET Scans

The earlier finding on univariate analysis of an increased proportion of PET scans being done in patients for whom a SOZ was not identified is perhaps an indicator of the fact that PET scans are reserved for the more complex cases. As these are thought to be particularly useful in lesion negative cases, we thought it worthwhile to explore this group further.

Of the 38 lesion negative cases, 35 underwent PET scans. Of these 35, a definable SOZ was identified through SEEG in 29 (82.9%). The localisation of the PET scan hypometabolism was compared to the localisation of the SOZ at the sub-lobar level from the text data on the data collection proforma. It was concordant in 14 (40.0%), falsely localising in 15 (42.9%), had wide PET abnormalities in 4 (11.4%) and was normal in 2 (5.7%). Twenty-two went on to have surgical treatment with 14 (63.6%) achieving an Engel class I outcome. The distribution by concordance is shown in Figure 4b.

## Discussion

We report a large multicentre retrospective series of 135 children with difficult-to-localise drug-resistant focal epilepsy undergoing 139 SEEG explorations. Overall, 86.7% of patients had a definable SOZ identified, 74.1% received subsequent surgical treatment and 34.8% had an Engel class I outcome at median follow-up of 1.3 years (Figure 5a). The Engel class I outcome in those undergoing surgical treatment was 47.0%, a figure comparable to other large single centre series and highlighting the complexity of the localisation of epilepsy in children undergoing SEEG. ^4,9,17^ Interestingly, this figure is slightly lower than the large series from Milan showing 59.4% ILAE class I-II outcomes (comparable to Engel class I) in a largely adult population, perhaps a reflection of the complex developmental and genetic aetiologies of the paediatric drug-resistant epilepsy population.^17^ Our cohort from 6 of the 7 UK paediatric SEEG centres adds a ‘real world’ perspective to the existing data as it represents the vast majority of UK paediatric SEEG cases to date and is representative of a national paediatric complex epilepsy population.

**Figure 5.**
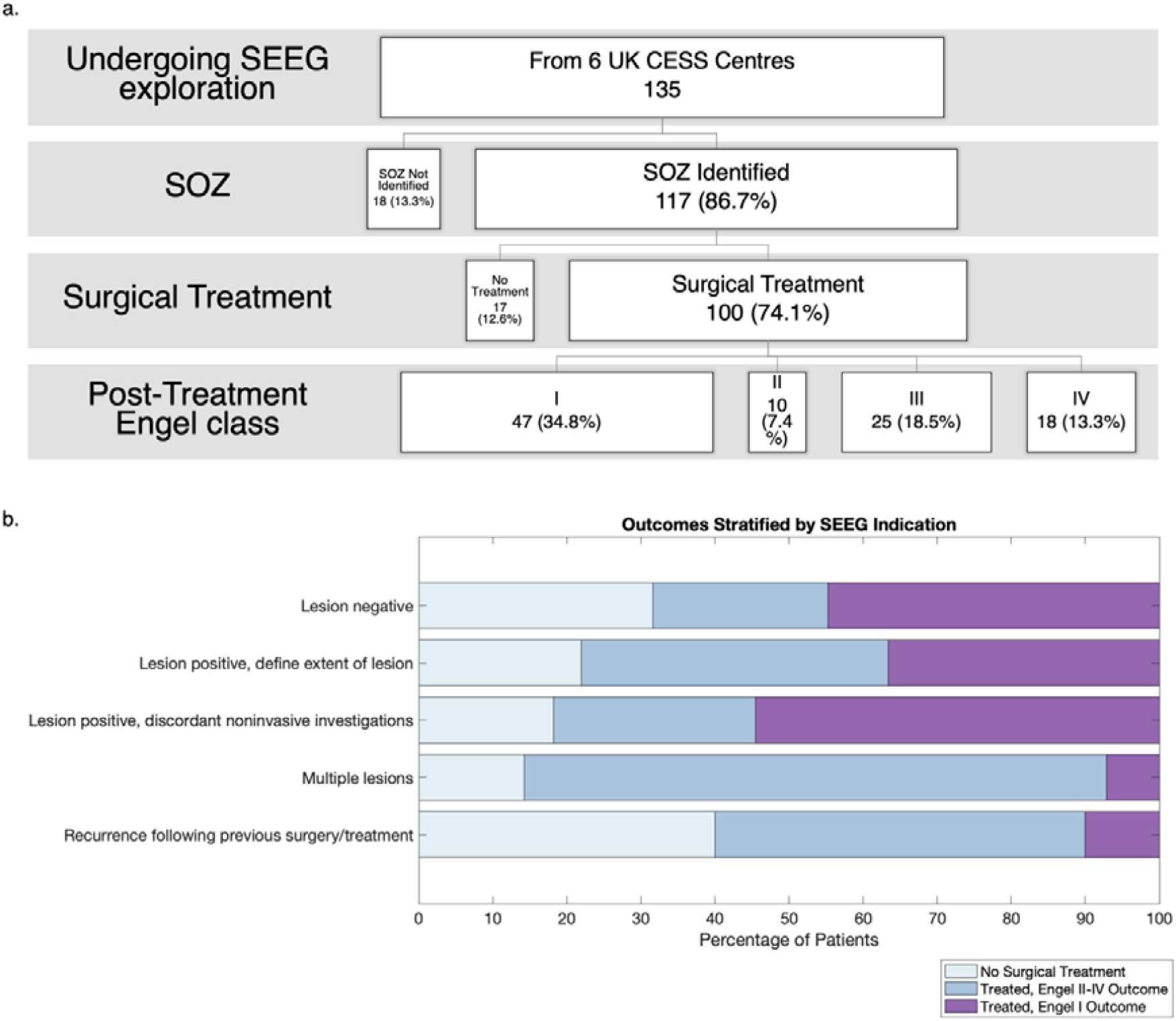
(a) Summary flowchart of outcomes in the 135 patients undergoing SEEG in this multi-centre retrospective UK cohort study. (b) Outcome of treatment stratified by indication for SEEG

A philosophical consideration that arises from these results surrounds the optimal target proportion of patients that should have a definable SOZ identified and have favourable outcomes following SEEG. This is dependent on patient selection thresholds, implantation strategy and subsequent interpretation of the SEEG findings (Figure 1). The proportions in this study represent a fair balance, where the majority (but not all) of those that are explored have a SOZ identified (86.7%) and those that go on to have surgical treatment, due to the inherent complexity, are less likely to achieve an Engel class I outcome (47.0%) than the more straightforward cases that do not require invasive intracranial evaluation. Other factors that could affect these proportions include the delineation of extent of the SOZ and subsequent surgical success of resecting this intended SOZ.

Two in-depth analyses were conducted, the first to identify factors associated with the identification of a SOZ and the second to identify factors associated with an Engel class I outcome following surgical treatment.

In the first analysis, the odds of successfully finding a definable SOZ was 3.6x more if ≥ 4 seizures were recorded compared to if <4 seizures were recorded during SEEG (p=0.03). This may provide increased confidence in a stereotyped pattern of seizures with onset in the same area. However, ≥ 4 seizures were recorded more commonly in those that did not become seizure free following surgery (Supplementary Table 4, p=0.07), indicating that factors that may improve the chances of SOZ identification may not necessarily be the same as those that improve chances of seizure freedom.

In the univariate analyses, the majority of significant factors included those directly related to the seizures, such as the number of seizures recorded, whether or not a seizure was stimulated and whether this stimulated seizure was a habitual seizure (Supplementary Table 3) all of which underscore the importance selecting patients that have frequent habitual seizures and stimulating these during intracranial recording.^18^

Another finding was that the odds of identifying a SOZ was 6.4x more for non-motor seizures, compared to motor seizures (p=0.03), a finding that has not been previously reported. Whilst there have been reports of high proportions of non-motor seizures in non-lesional epilepsy cohorts, this was not the case in our cohort.^19^ This perhaps reflects the difficulty in children of ascertaining accurate non-motor semiology; many of the cases classified as motor semiology may in fact have preceding non-motor manifestations that were not able to be described accurately by the children. A more detailed analysis of the scalp EEG video-telemetry results might shed light as to whether there were electrographic changes prior to motor onset suggesting that a non-motor onset may have been missed.

In the second analysis, the only significant factor associated with an Engel class I outcome was indication for SEEG (p=0.03). The odds of a class I outcome were 5.9x less with an indication of recurrence following previous surgical treatment as compared to the lesion negative indication (p=0.002). Interestingly, when viewed as a function of all explorations, both the recurrence and multiple lesion cohorts have much poorer overall outcomes compared to the other indications (Figure 5b). As a result of this finding, we explored the recurrence and TSC (vast majority of the multiple lesions) cohorts further. In the recurrence cohort, only 10.0% went on to become seizure free. None of the repeat lesionectomy patients (including 5 with FCD IIa or IIb histology) became seizure free, although we were not aware of whether these were focal resections adjacent to (ie residual lesion) or distant from the original resections.^20^ Irrespective of this, the finding reinforces the concept of ‘surgical refractoriness’ that has been recently purported in the literature.^21^ It suggests that those with ongoing seizures or recrudescence following surgery probably warrant consideration of more aggressive approaches (such as larger lobar resections, TPO disconnection or hemispherotomy), although the risks and benefits need to be assessed on an individual bases. Quantitative analyses in these patients may also be a helpful adjunct to assess how the network architecture changes to support ongoing seizures following initial surgery.^22^

Although not significant (p=0.10) on the final regression analysis, the histology had an important bearing on the outcome following SEEG-guided surgical treatment. Consistent with the established literature, those with a diagnosis of FCD Type IIa or IIb had a 8.9x and 10.4x higher odds of seizure freedom (p=0.02 and p=0.01 respectively) compared to a non-diagnostic/other histology.^23^ However, histology and indication for SEEG covaried in a way that histology lost significance in the final analysis.

Despite complex resections involving multiple tubers and mesial temporal structures, outcomes were poor in the cohort with TSC that were explored with SEEG, with only 7.7% achieving and Engel class I outcome. However, a total of 69.2% achieved class I-III outcomes, indicating at least a worthwhile improvement following epilepsy surgery; in some of these cases, patients would have had multiple seizure types with the explicit understanding to target only one (eg the most disabling) during SEEG and subsequent resective surgery. This highlights the complexity of the epileptogenic networks in TSC. A recent national series from China, where they performed a combination of tuber-only, tuber + surrounding cortex and larger lobar resections has demonstrated that good outcomes are possible in tuberous sclerosis with over 70% achieving seizure freedom at 1 year and 60% at 4 years.^24^ Going forwards, comparisons need to be made to outcomes in children undergoing resection in TSC without SEEG to assess whether these poor outcomes are restricted to the small number of more complex patients. This will allow a critical view on whether certain factors (eg tuber burden, presence of single large/outstanding tuber, presence of multiple semiologies, EEG characteristics) predict for poor outcome in TSC, which will help refine the choice of candidates for SEEG exploration.

The utility of PET in MRI lesion negative patients remains an area of interest and has been found to be useful in patients with malformations of cortical development undergoing SEEG. ^25,26^ In this study, the PET analysis was limited to text fields in the data collection proforma and the imaging was not formally reviewed; therefore, the findings must be interpreted with caution. Given the limitations, we found that PET hypometabolism was not concordant with the SEEG-defined SOZ in 60.0% of cases. When those in whom an SOZ was not identified are removed, the proportion of patients who an Engel Class I outcome was similar irrespective of PET findings (Figure 4b). What this study was not able to establish was the added value of the PET scan – although it was concordant in 40.0% of cases, it is arguable that it could be misleading in the remaining 60.0%; the impact of PET information (and for that matter other adjunctive investigations such as MEG and ictal SPECT) on the hypothesis generation and planning of SEEG electrode locations is difficult to assess retrospectively and requires careful prospective study designs to ascertain the true impact. However, a retrospective finding of concordance (or not) with the SEEG-defined SOZ could help in counselling patients/parents about the chances of seizure freedom following resective surgery.

### Limitations

Surgical failures are presumed due to either inaccurate localisation or incomplete resection of the epileptogenic zone.^4^ Although many localisation factors have been considered in this study, we did not analyse specific features of the SEEG recordings at seizure onset. Previous studies have shown that certain pathologies may associated with specific patterns of EEG change at seizure onset, some of which may be associated with better post-surgical outcomes.^27–29^ Instead, we used expert neurophysiologist-reported assessment as a measure of seizure onset. Another limitation of our study is that we did not assess the completeness of resection of the SOZ.^17^ In these difficult-to-localise cases, this is often challenging to assess as it is not limited to just the MRI-visible lesion and requires additional post-operative image post-processing to specifically identify which contacts have been resected. Despite these limitations, we have considered a comprehensive list of factors from the non-invasive evaluation and the SEEG procedure that shed light on this complex population of children. We envisage that the results will be useful to multidisciplinary teams planning SEEG in children and in the counselling of children and families prior to undertaking SEEG.

In addition, the study is susceptible to all the traditional biases of a multi-centre retrospective cohort study. The rarity and complexity of SEEG ensures that cases were not missed in this cohort, although there remains recall bias associated with gathering data retrospectively from clinical records. This cohort does not consider the alternative intracranial evaluation to SEEG, which is subdural electrodes. Given the low morbidity and numerous advantages of SEEG, the current use of grids and strips is relatively limited within the UK CESS network and the nuances of selection between these two types of intracranial EEG evaluation remain to be fully elucidated.^30^

### Future directions

In addition to refining the selection of patients, implantation strategy and subsequent surgical planning in SEEG patients using clinical data, we are likely to see increasing incorporation of quantitative methods in SEEG planning, including automated analysis of MRI,^31^ and computational analysis of SEEG recordings.^22,32,33^ Whilst seizure (both spontaneously recorded and stimulated) have been shown to be crucial to outcomes in this present series, concepts such as identification of the SOZ from interictal recordings^34^, using additional methods such as microelectrode recordings^35^ or network-based analyses^22^ may improve the interpretation of SEEG recordings as we move from a location-focused to network-based interventions.

## Conclusion

In this large multi-centre series of 135 children undergoing 139 SEEG explorations in the UK, we demonstrate that 86.7% of patients had a definable SOZ identified, 74.1% received subsequent surgical treatment and 34.8% had an Engel class I at a median follow-up of 1.3 years. Of those undergoing SEEG-guided surgical treatment, 47.0% achieved an Engel class I outcome. Seizure semiology and number of seizures recorded were important factors associated with the identification of a definable SOZ whilst indication for SEEG was the most important factor associated with post-surgical outcome.

We also demonstrate that SEEG is safe with only one (0.7%) haematoma requiring surgical evacuation and no procedure-related long-term neurological deficits. Epilepsy in children that requires intracranial evaluation prior to surgical intervention is a complex entity and this study highlights the positive impact that can be had as a result of SEEG exploration in this cohort as 84.0% of those undergoing SEEG-guided surgical treatments experience at least a worthwhile improvement with 47.0% achieving seizure freedom.

## Data Availability

Individual patient-level data, including demographics, non-invasive evaluation, SEEG factors, operative factors and outcome measures are available from the corresponding author on request. The final decision to share the data remains at the discretion of the corresponding author and will be based on approval of a proposal for the use of the data. 

## Children’s Epilepsy Surgery Service Collaborators List

### Great Ormond Street Hospital, London

Aswin Chari

Friederike Moeller

Stewart Boyd

M Zubair Tahir

J Helen Cross

Christin Eltze

Krishna Das

Thijs van Dalen

Rod C Scott

Ronit Pressler

Rachel C Thornton

Martin M Tisdall

### Bristol Royal Hospital for Children, Bristol

Elliott Warren

Jayesh Patel

Michael Carter

Nicholas Kane

Andrew A Mallick

Marcus Likeman

Sarah Rushton

Danielle Cole

### Manchester Children’s Hospital, Manchester

Athi Ponnusamy

Jeen Tan

Jonathan Ellenbogen

John Kitchen

Majid Aziz

Stuart Rust

### Alder Hey Children’s Hospital, Liverpool

Sona Janackova

Sasha Burn

Anand Iyer

### Royal Hospital for Sick Children, Edinburgh

Jay Shetty

Ailsa McLellan

Jothy Kandasamy

Drahoslav Sokol

### Kings Health Partners (Kings College Hospital & Evelina London Children’s Hospital), London

Elaine Hughes

Harutomo Hasegawa

Richard Selway

Harishchandra Srinivasan

Rinki Singh

Nandini Mullatti

Franz Brunnhuber

Zaloa Agirre-Arrizubieta

Robert Elwes

Sushma Goyal

Antonio Valentin

Ioannis Stavropoulos

## Supplementary Material

STROBE Statement—Checklist of items that should be included in reports of ***cohort studies***

|  | Item No | Recommendation | Page No |
| --- | --- | --- | --- |
| **Title and abstract** | 1 | (a) Indicate the study's design with a commonly used term in the title or the abstract | 1 |
|  |  | (b) Provide in the abstract an informative and balanced summary of what was done and what was found | 2 |
| **Introduction** |  |  |  |
| Background/rationale | 2 | Explain the scientific background and rationale for the investigation being reported | 4 |
| Objectives | 3 | State specific objectives, including any prespecified hypotheses | 4 |
| **Methods** |  |  |  |
| Study design | 4 | Present key elements of study design early in the paper | 5 |
| Setting | 5 | Describe the setting, locations, and relevant dates, including periods of recruitment, exposure, follow-up, and data collection | 5 |
| Participants | 6 | (a) Give the eligibility criteria, and the sources and methods of selection of participants. Describe methods of follow-up<br>(b) For matched studies, give matching criteria and number of exposed and unexposed | 5 |
| Variables | 7 | Clearly define all outcomes, exposures, predictors, potential confounders, and effect modifiers. Give diagnostic criteria, if applicable | 5 |
| Data sources/<br>measurement | 8* | For each variable of interest, give sources of data and details of methods of assessment (measurement). Describe comparability of assessment methods if there is more than one group | 5 |
| Bias | 9 | Describe any efforts to address potential sources of bias | 5 |
| Study size | 10 | Explain how the study size was arrived at | N/A |
| Quantitative variables | 11 | Explain how quantitative variables were handled in the analyses. If applicable, describe which groupings were chosen and why | 6 |
| Statistical methods | 12 | (a) Describe all statistical methods, including those used to control for confounding<br>(b) Describe any methods used to examine subgroups and interactions<br>(c) Explain how missing data were addressed<br>(d) If applicable, explain how loss to follow-up was addressed<br>(e) Describe any sensitivity analyses | 6 |
| **Results** |  |  |  |
| Participants | 13* | (a) Report numbers of individuals at each stage of study—eg numbers potentially eligible, examined for eligibility, confirmed eligible, included in the study, completing follow-up, and analysed<br>(b) Give reasons for non-participation at each stage<br>(c) Consider use of a flow diagram | 7 |
| Descriptive data | 14* | (a) Give characteristics of study participants (eg demographic, clinical, social) and information on exposures and potential confounders<br>(b) Indicate number of participants with missing data for each variable of interest<br>(c) Summarise follow-up time (eg, average and total amount) | 7 |
| Outcome data | 15* | Report numbers of outcome events or summary measures over time | 8 |
| Main results | 16 | (a) Give unadjusted estimates and, if applicable, confounder-adjusted estimates and their precision (eg, 95% confidence interval). Make clear which confounders were adjusted for and why they were included<br>(b) Report category boundaries when continuous variables were categorized<br>(c) If relevant, consider translating estimates of relative risk into absolute risk for a meaningful time period | 8 |
| Other analyses | 17 | Report other analyses done—eg analyses of subgroups and interactions, and sensitivity analyses | 9 |
| **Discussion** |  |  |  |
| Key results | 18 | Summarise key results with reference to study objectives | 11 |
| Limitations | 19 | Discuss limitations of the study, taking into account sources of potential bias or imprecision. Discuss both direction and magnitude of any potential bias | 13 |
| Interpretation | 20 | Give a cautious overall interpretation of results considering objectives, limitations, multiplicity of analyses, results from similar studies, and other relevant evidence | 14 |
| Generalisability | 21 | Discuss the generalisability (external validity) of the study results | 14 |
| **Other information** |  |  |  |
| Funding | 22 | Give the source of funding and the role of the funders for the present study and, if applicable, for the original study on which the present article is based | 6 |

\*Give information separately for exposed and unexposed groups.
**Note:** An Explanation and Elaboration article discusses each checklist item and gives methodological background and published examples of transparent reporting. The STROBE checklist is best used in conjunction with this article (freely available on the Web sites of PLoS Medicine at http://www.plosmedicine.org/, Annals of Internal Medicine at http://www.annals.org/, and Epidemiology at http://www.epidem.com/). Information on the STROBE Initiative is available at http://www.strobe-statement.org.

**Table 1:** Domains of data collection proforma used in this multicentre study. ILAE = International League Against Epilepsy, ASM = antiseizure medication, MRI = magnetic resonance imaging, EEG = electroencephalography, LITT = laser interstitial thermal therapy, SOZ = seizure onset zone, TSC = tuberous sclerosis complex

| Domain | Example Data Points |
| --- | --- |
| Demographic & Seizure History | Age, age of seizure onset, seizure type(s) according to the 2017 ILAE operational classification, number of previous and current ASMs, previous surgical intervention |
| Non-invasive evaluation | MRI abnormality and location, EEG video-telemetry ictal and interictal lateralisation/localisation, other non-invasive investigations |
| Indication for SEEG | Catergorised as below:<br>- Lesion negative: No or subtle/non-specific MRI abnormality<br>- Lesion positive, discordant non-invasive investigations: EEG, semiology or PET disagreeing with MRI lesion localisation<br>- Lesion positive, define extent of lesion: Widespread lesion or close to eloquent cortex that required definition of its extent and/or functional tissue<br>- Multiple lesions: Most commonly, due to TSC<br>- Recurrence following previous surgery/treatment: Previous surgical treatment for epilepsy including resective & disconnective surgery. |
| SEEG factors | Number of electrodes, number of lobes, duration of recording, seizures recorded, seizures stimulated? |
| Operative factors | Type of surgical intervention (lesionectomy, lobectomy, thermocoagulation, LITT, hemispherotomy or disconnection), |
| Outcome measures | (a) Identification of a definable SOZ<br>(b) Post-operative seizure outcome (Engel class) at last follow-up following surgical resection |

**Table 2:**
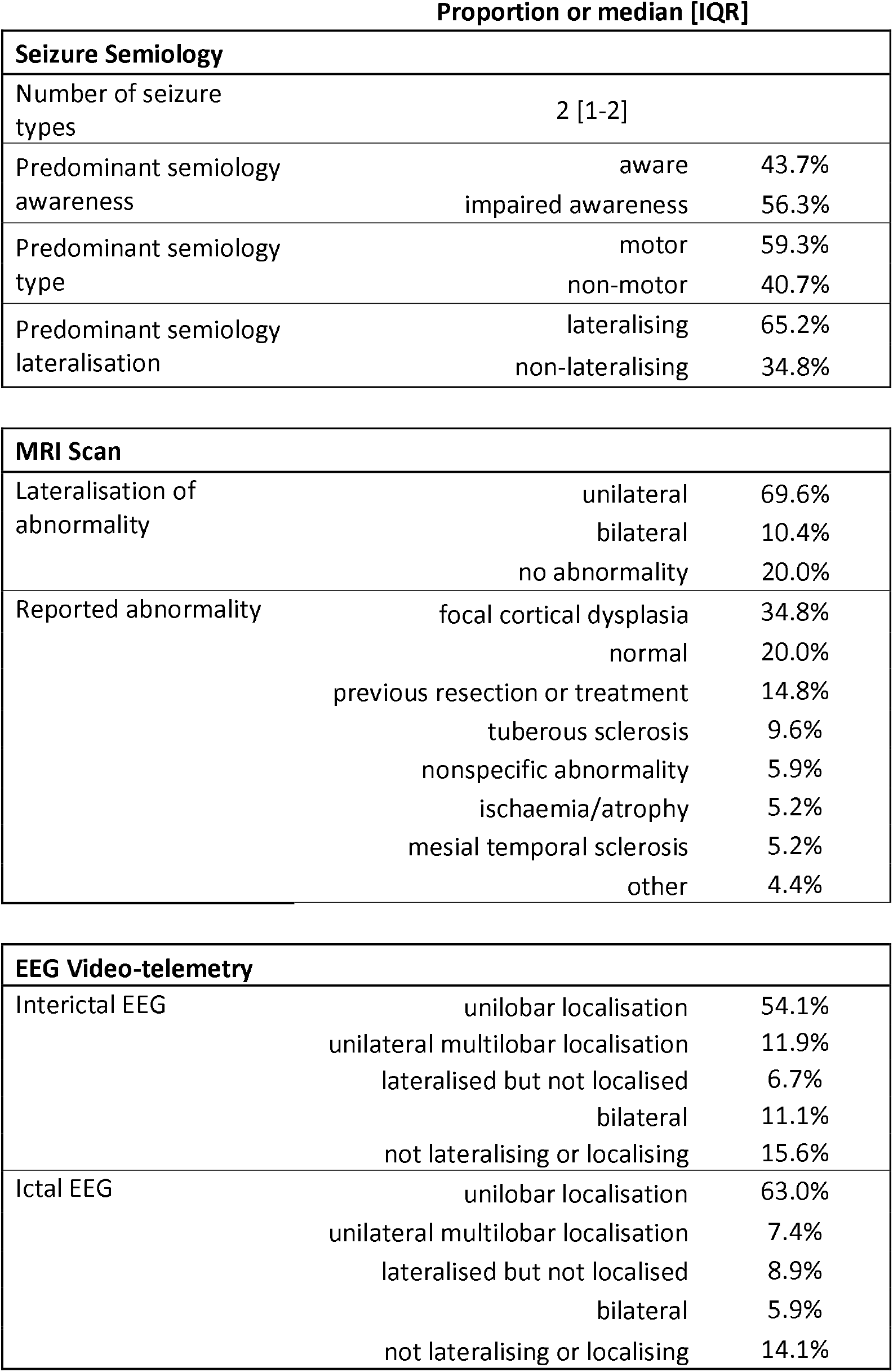
Details of the non-invasive evaluation, outlining the proportions of patients in each category (for categorical variables) or the median [IQR] (for discrete variables).

**Table 3:**
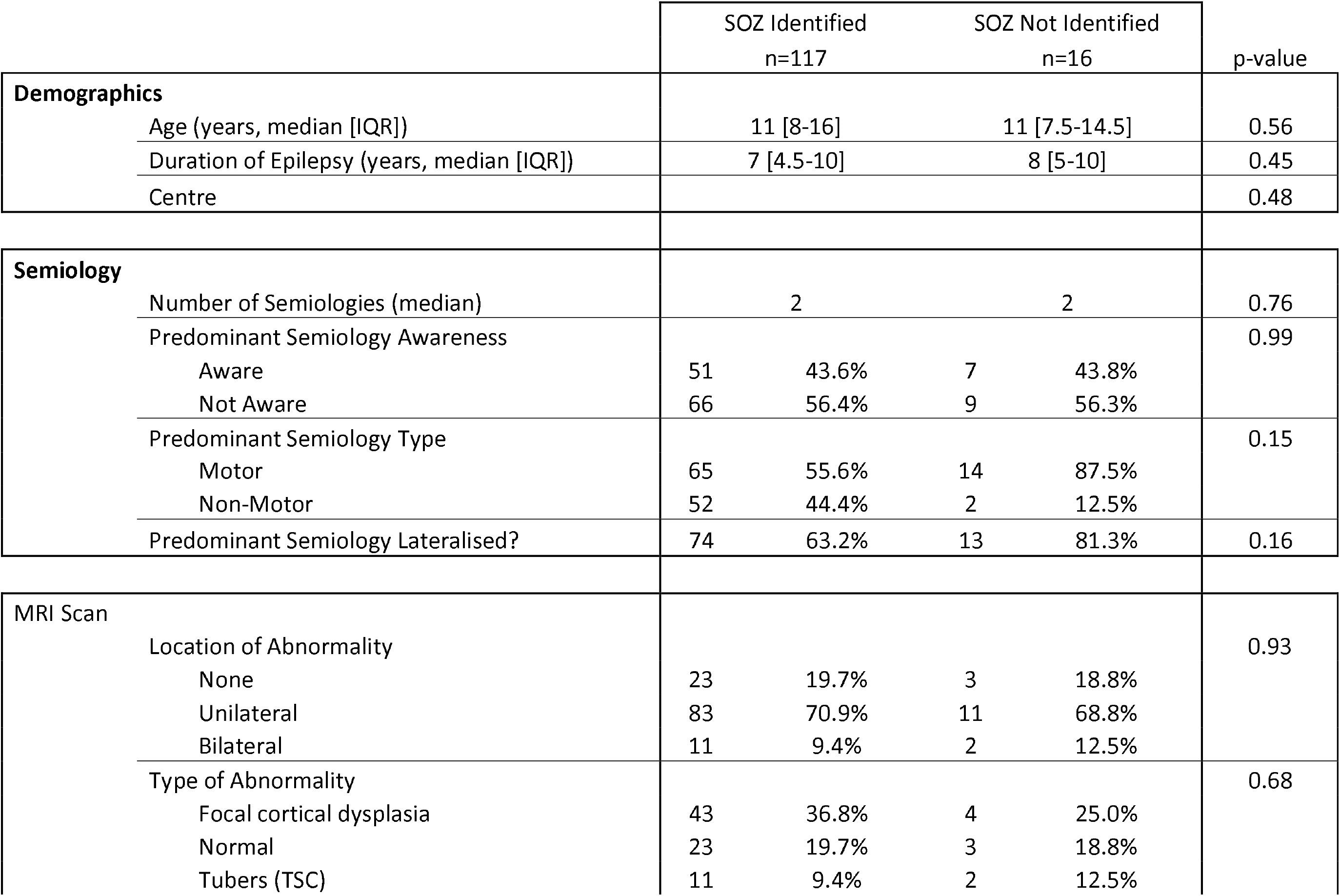
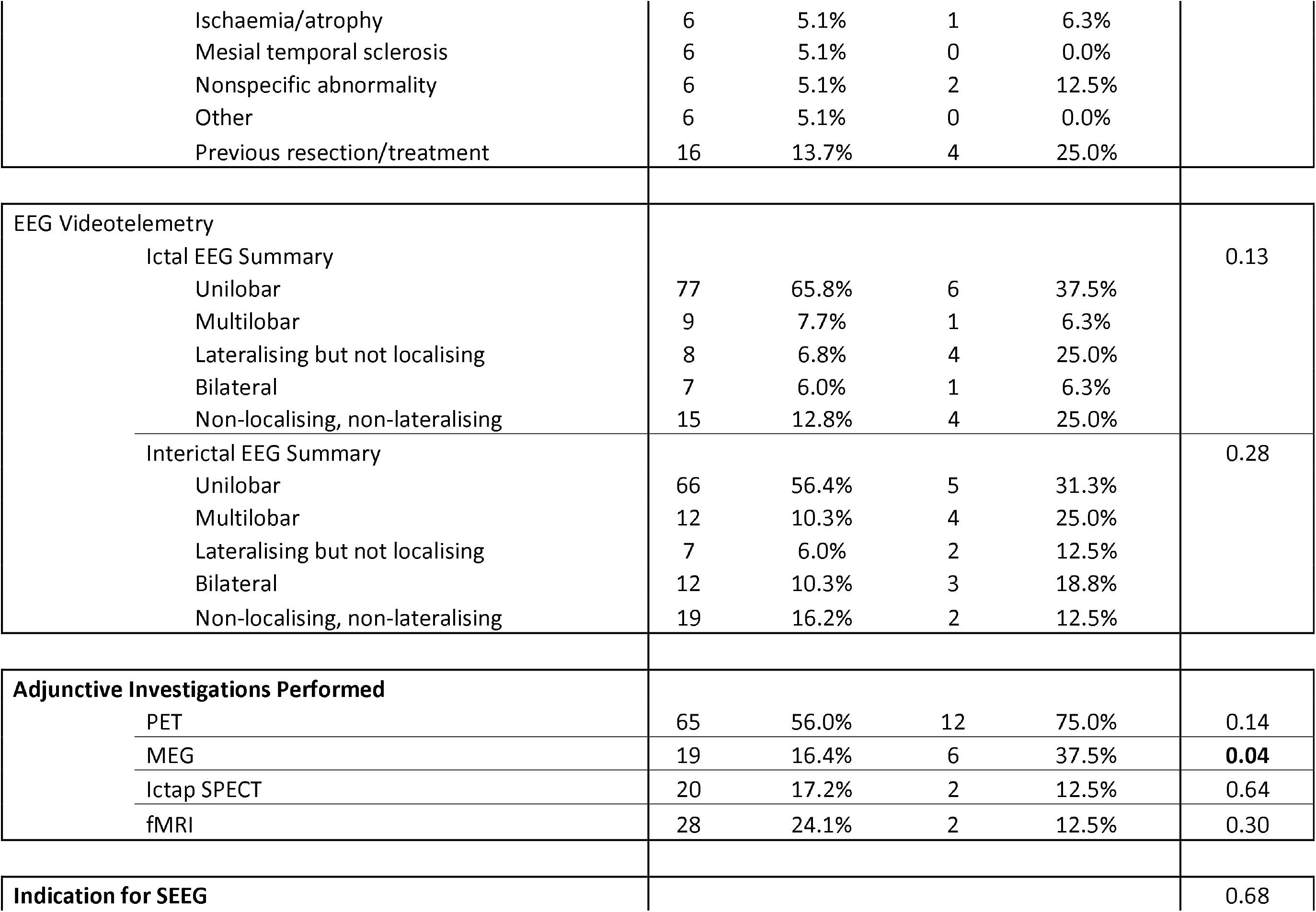
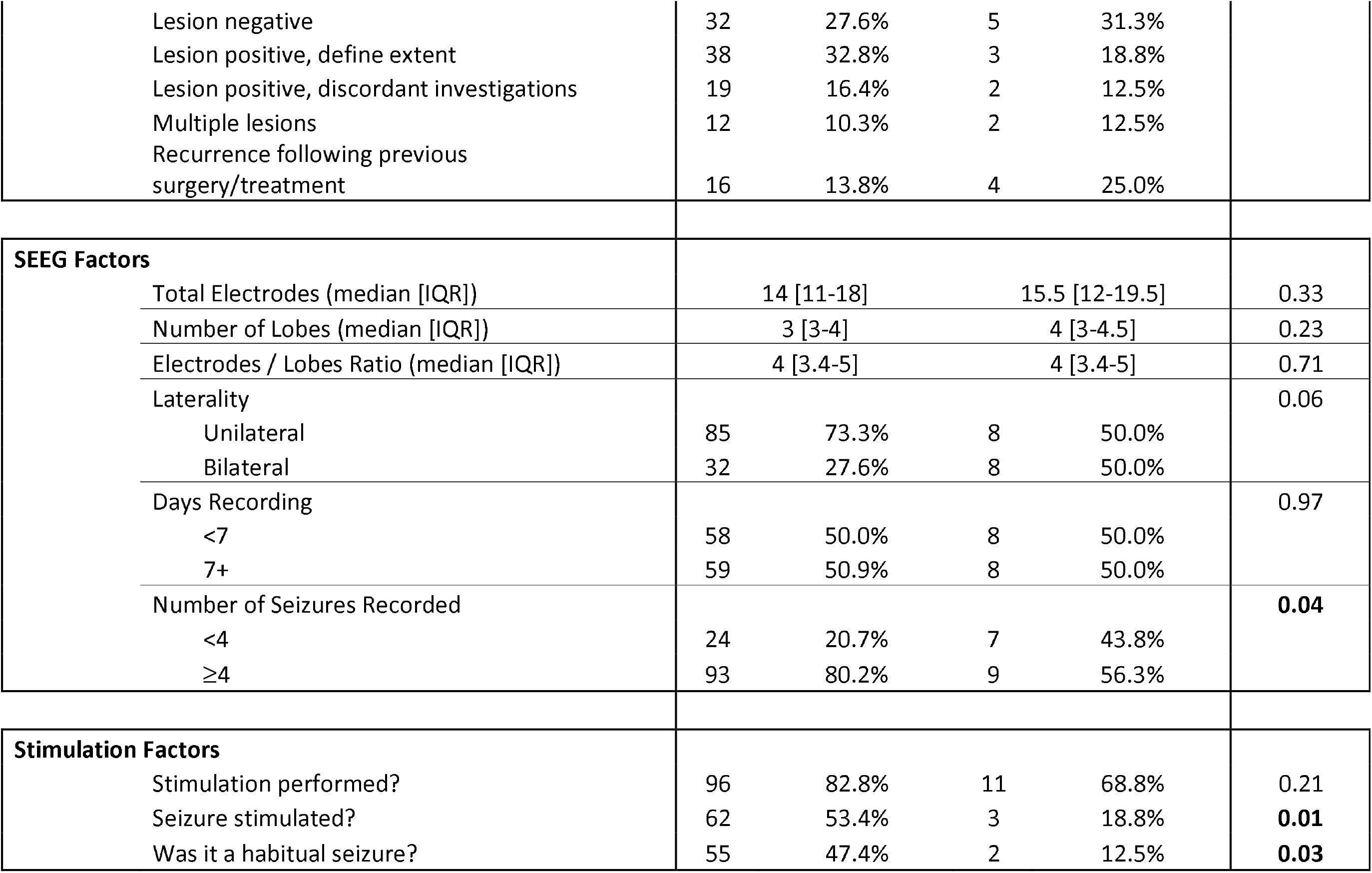
Factors associated with whether or not a SOZ was identified on SEEG in 133 patients undergoing SEEG. Comparisons were made using Kruskal-Wallis tests for continuous variables and chi-squared tests for categorical variables.

**Table 4:**
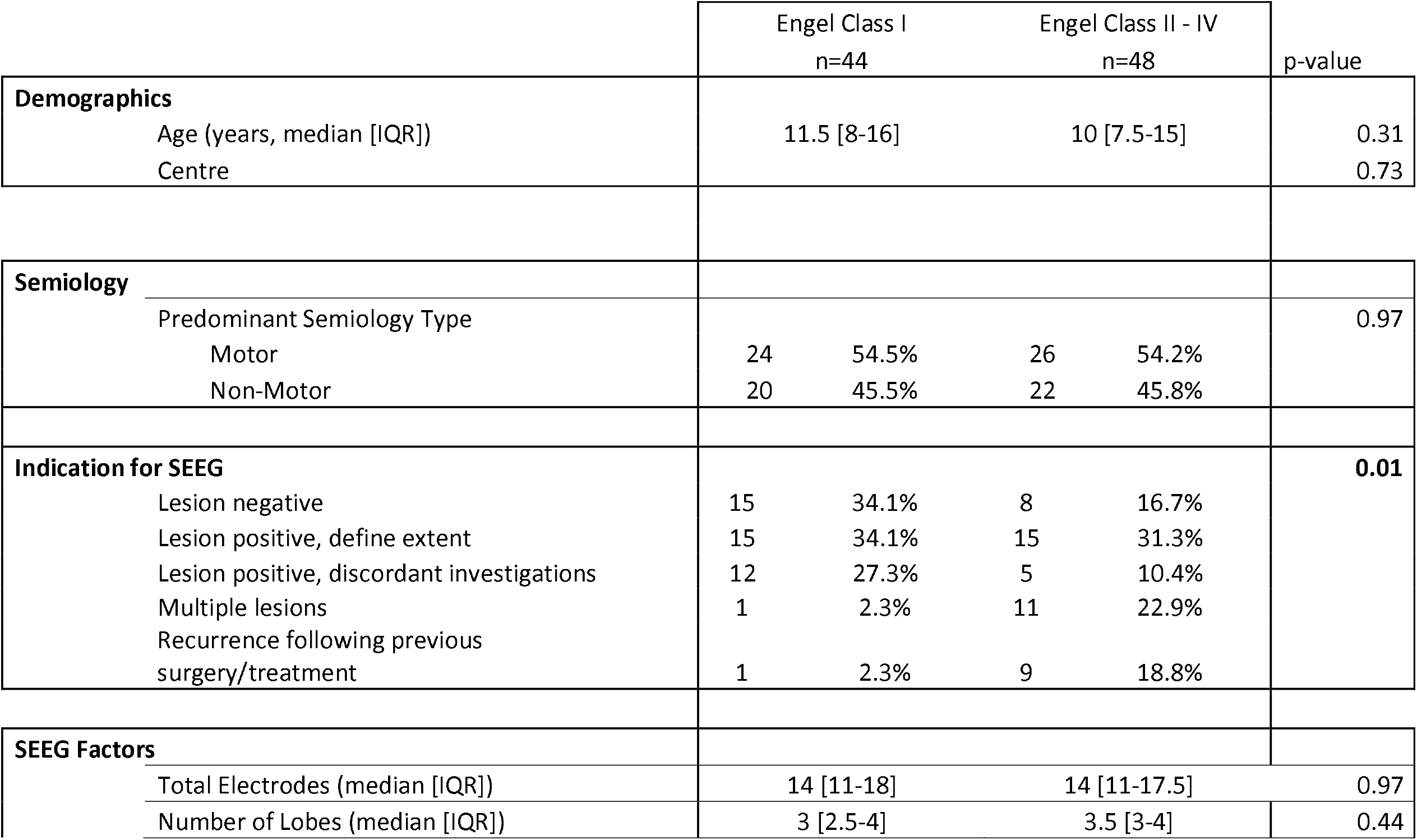
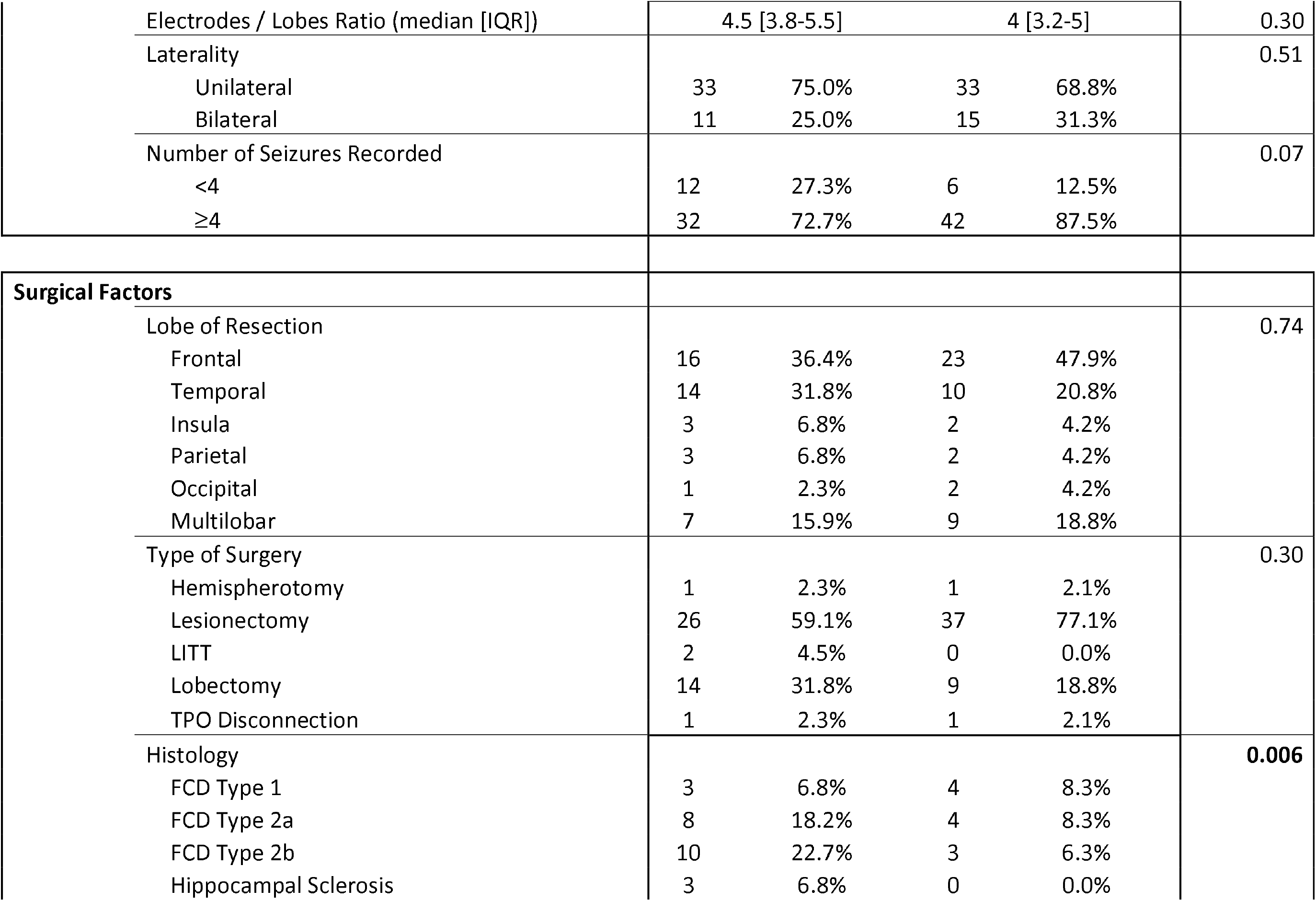
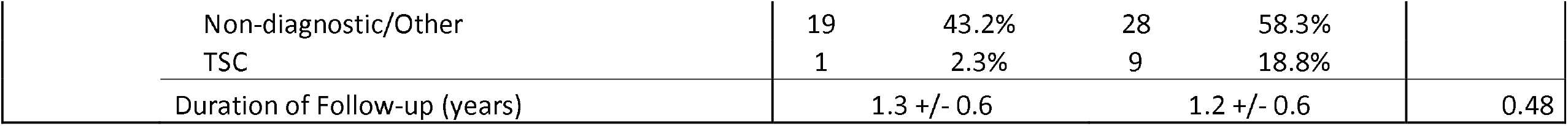
Factors associated with favourable or unfavourable outcome in 92 patients undergoing SEEG-guided tailored treatments. LITT = laser interstitial thermal therapy, TPO = temporo-parieto-occipital, FCD = focal cortical dysplasia, TSC = tuberous sclerosis complex

## References

1 Barba C, Cross JH, Braun K, et al. Trends in pediatric epilepsy surgery in Europe between 2008 and 2015: Country-, center-, and age-specific variation. Epilepsia 2019; published online Dec 26. DOI:10.1111/epi.16414.

2 Braun KPJ. Influence of epilepsy surgery on developmental outcomes in children. European Journal of Paediatric Neurology 2020; 24:40–2.

3 Taussig D, Chipaux M, Fohlen M, et al. Invasive evaluation in children (SEEG vs subdural grids). Seizure 2018; published online Nov 16. DOI:10.1016/j.seizure.2018.11.008.

4 McGovern RA, Knight EP, Gupta A, et al. Robot-assisted stereoelectroencephalography in children. J Neurosurg Pediatr 2018; 23:288–96.

5 Abel TJ, Varela Osorio R, Amorim-Leite R, et al. Frameless robot-assisted stereoelectroencephalography in children: technical aspects and comparison with Talairach frame technique. J Neurosurg Pediatr 2018; 22:37–46.

6 Ho AL, Muftuoglu Y, Pendharkar AV, et al. Robot-guided pediatric stereoelectroencephalography: single-institution experience. J Neurosurg Pediatr 2018; 22:1–8.

7 Ho AL, Feng AY, Kim LH, et al. Stereoelectroencephalography in children: a review. Neurosurg Focus 2018; 45:E7.

8 Sharma JD, Seunarine KK, Tahir MZ, Tisdall MM. Accuracy of robot-assisted versus optical frameless navigated stereoelectroencephalography electrode placement in children. J Neurosurg Pediatr 2019; 23:297–302.

9 Taussig D, Chipaux M, Lebas A, et al. Stereo-electroencephalography (SEEG) in 65 children: an effective and safe diagnostic method for pre-surgical diagnosis, independent of age. Epileptic Disord 2014; 16:280–95.

10 Dorfmüller G, Ferrand-Sorbets S, Fohlen M, et al. Outcome of surgery in children with focal cortical dysplasia younger than 5 years explored by stereo-electroencephalography. Childs Nerv Syst 2014; 30:1875–83.

11 Budke M, Avecillas-Chasin JM, Villarejo F. Implantation of Depth Electrodes in Children Using VarioGuide® Frameless Navigation System: Technical Note. Oper Neurosurg (Hagerstown) 2018; 15:302–9.

12 Goldstein HE, Youngerman BE, Shao B, et al. Safety and efficacy of stereoelectroencephalography in pediatric focal epilepsy: a single-center experience. J Neurosurg Pediatr 2018; 22:444–52.

13 Sacino MF, Huang SS, Schreiber J, Gaillard WD, Oluigbo CO. Is the use of Stereotactic Electroencephalography Safe and Effective in Children? A Meta-Analysis of the use of Stereotactic Electroencephalography in Comparison to Subdural Grids for Invasive Epilepsy Monitoring in Pediatric Subjects. Neurosurgery 2018; published online Oct 22. DOI:10.1093/neuros/nyy466.

14 Tomlinson SB, Buch VP, Armstrong D, Kennedy BC. Stereoelectroencephalography in Pediatric Epilepsy Surgery. J Korean Neurosurg Soc 2019; 62:302–12.

15 Liu Y, Chen G, Chen J, et al. Individualized stereoelectroencephalography evaluation and navigated resection in medically refractory pediatric epilepsy. Epilepsy Behav 2020; 112:107398.

16 von Elm E, Altman DG, Egger M, et al. The Strengthening the Reporting of Observational Studies in Epidemiology (STROBE) statement: guidelines for reporting observational studies. J Clin Epidemiol 2008; 61:344–9.

17 Cardinale F, Rizzi M, Vignati E, et al. Stereoelectroencephalography: retrospective analysis of 742 procedures in a single centre. Brain 2019; 142:2688–704.

18 George DD, Ojemann SG, Drees C, Thompson JA. Stimulation Mapping Using Stereoelectroencephalography: Current and Future Directions. Front Neurol 2020; 11:320.

19 Süße M, Hamann L, Flöel A, Podewils F von. Nonlesional late-onset epilepsy: Semiology, EEG, cerebrospinal fluid, and seizure outcome characteristics. Epilepsy Behav 2019; 91:75–80.

20 Vaugier L, Lagarde S, McGonigal A, et al. The role of stereoelectroencephalography (SEEG) in reevaluation of epilepsy surgery failures. Epilepsy Behav 2018; 81:86–93.

21 Yardi R, Morita-Sherman ME, Fitzgerald Z, et al. Long-term outcomes of reoperations in epilepsy surgery. Epilepsia 2020; 61:465–78.

22 Bartolomei F, Lagarde S, Wendling F, et al. Defining epileptogenic networks: Contribution of SEEG and signal analysis. Epilepsia 2017; 58:1131–47.

23 Blumcke I, Spreafico R, Haaker G, et al. Histopathological Findings in Brain Tissue Obtained during Epilepsy Surgery. New England Journal of Medicine 2017; 377:1648–56.

24 Liu S, Yu T, Guan Y, et al. Resective epilepsy surgery in tuberous sclerosis complex: a nationwide multicentre retrospective study from China. Brain 2020; 143:570–81.

25 Lagarde S, Boucekine M, McGonigal A, et al. Relationship between PET metabolism and SEEG epileptogenicity in focal lesional epilepsy. Eur J Nucl Med Mol Imaging 2020; published online May 20. DOI:10.1007/s00259-020-04791-1.

26 Hu W-H, Wang X, Liu L-N, et al. Multimodality Image Post-processing in Detection of Extratemporal MRI-Negative Cortical Dysplasia. Front Neurol 2018; 9:450.

27 Lagarde S, Buzori S, Trebuchon A, et al. The repertoire of seizure onset patterns in human focal epilepsies: Determinants and prognostic values. Epilepsia 2019; 60:85–95.

28 Di Giacomo R, Uribe-San-Martin R, Mai R, et al. Stereo-EEG ictal/interictal patterns and underlying pathologies. Seizure 2019; 72:54–60.

29 Jiménez-Jiménez D, Nekkare R, Flores L, et al. Prognostic value of intracranial seizure onset patterns for surgical outcome of the treatment of epilepsy. Clin Neurophysiol 2015; 126:257–67.

30 Englot DJ. Surface or Depth: A Paradigm Shift in Invasive Epilepsy Monitoring. Epilepsy Curr 2020; : 1535759720949248.

31 Wagstyl K, Adler S, Pimpel B, et al. Planning stereoelectroencephalography using automated lesion detection: Retrospective feasibility study. Epilepsia 2020; 61:1406–16.

32 Balatskaya A, Roehri N, Lagarde S, et al. The “Connectivity Epileptogenicity Index “ (cEI), a method for mapping the different seizure onset patterns in StereoElectroEncephalography recorded seizures. Clin Neurophysiol 2020; 131:1947–55.

33 Andrzejak RG, David O, Gnatkovsky V, et al. Localization of Epileptogenic Zone on Pre-surgical Intracranial EEG Recordings: Toward a Validation of Quantitative Signal Analysis Approaches. Brain Topography 2015; 28:832–7.

34 Goodale SE, González HFJ, Johnson GW, et al. Resting-State SEEG May Help Localize Epileptogenic Brain Regions. Neurosurgery 2019; published online Sept 16. DOI:10.1093/neuros/nyz351.

35 Chari A, Thornton RC, Tisdall MM, Scott RC. Microelectrode recordings in human epilepsy: A case for clinical translation? Brain Commun 2020. DOI:10.1093/braincomms/fcaa082.

36 Kokkinos V, Richardson RM. Epilepsy Surgery: The Network Approach. Neurosurgery Clinics of North America 2020; 31:i.

